# HiSpike: A high-throughput cost effective sequencing method for the SARS-CoV-2 spike gene

**DOI:** 10.1101/2021.03.02.21252290

**Authors:** Ephraim Fass, Gal Zizelski Valenci, Mor Rubinstein, Paul J Freidlin, Shira Rosencwaig, Inna Kutikov, Robert Werner, Nofar Ben-Tovim, Efrat Bucris, Neta S Zuckerman, Orna Mor, Ella Mendelson, Zeev Dveyrin, Efrat Rorman, Israel Nissan

## Abstract

The changing nature of the corona virus of the SARS-CoV-2 pandemic poses unprecedented challenges to the world’s health systems. New and virulent emerging spike gene variants, such as the UK 20I/501Y.V1 and South African 20H/501Y.V2, could jeopardize global efforts to produce immunity and reduce mortality. These challenges require effective real-time genomic surveillance solutions that the medical community can quickly adopt. The SARS-CoV-2 spike protein mediates host receptor recognition and entry into the cell and therefore, it is most susceptible to generation of variants with increased transmissibility and pathogenicity. The spike protein is also the primary target of neutralizing antibodies in COVID-19 patients and the most common antigen for induction of effective vaccine immunity. Therefore, tight monitoring of the spike protein gene variants is key to mitigating COVID-19 spread and vaccine escape mutants. Currently, the ARTIC method for SARS-CoV-2 whole genome sequencing is applied worldwide. However, this method commonly requires more than 96 hours (4-5 days) from start to finish and at present high sample sequence demands, sequencing resources are quickly exhausted. In this work, we present HiSpike, a method for high-throughput targeted next generation sequencing (NGS) of the spike gene. This simple three-step method can be completed in less than 30 hours and can sequence 10-fold more samples compared to the conventional ARTIC method and at a fraction of the cost. HiSpike was proven valid, and has identified, at high quality, multiple spike variants from real-time field samples, such as the UK and the South African variants. This method will certainly be effective in discovering future spike mutations. Therefore, running HiSpike for full sequencing of the spike gene of all positive SARS-CoV-2 samples could be considered for near real-time detection of known and emerging spike mutations as they evolve. HiSpike provides affordable sequencing options to help laboratories conserve resources, hence it provides a tool for widespread monitoring, that can support critical knowledge-based decisions.

## INTRODUCTION

Mutations of SARS-CoV-2 spike glycoprotein changed COVID-19 from a local outbreak in Wuhan, China [1-4] into a worldwide pandemic [5-8], inflicting morbidity and mortality on currently over 100 million people with nearly 2.5 million deaths. COVID-19 has severely disrupted health and educational systems, and continues to wreak social, cultural, and economic havoc in afflicted countries.

The spike protein, a homotrimeric class I fusion glycoprotein, is responsible for the ability of the SARS-CoV-2 virus to recognize and bind to the host cell receptor angiotensin converting enzyme 2 (ACE2) [9-14], and to induce subsequent membrane fusion and viral entry into the host cell [15, 16]. It is thought that mutation-driven changes to the spike protein promoted the initial zoonotic event of coronavirus jump from animal reservoir to human [17-19], and that continued mutation-driven change of SARS-CoV-2 spike protein in COVID-19 patients promotes increasing adaptation of the virus to the human host [20-22]. Indeed, mutations in SARS-CoV-2 spike glycoprotein have resulted in vastly increased transmissibility, infectivity and viral load in humans [6, 7, 23-25].

Now, a year into the pandemic, we observe multiple new, predominantly spike protein variants, with superior transmissibility that are changing the dynamics of the pandemic. Currently leading in prevalence, in Europe and Israel, are the spike mutation variants UK 20I/501Y.V1 (also known as B.1.1.7) and South African 20H/501Y.V2 (also known as B.1.351) [26-28] (https://virological.org/t/mutations-arising-in-sars-cov-2-spike-on-sustained-human-to-human-transmission-and-human-to-animal-passage/578). The UK variant has eight non-synonymous mutations and deletions located in the spike gene (https://virological.org/t/preliminary-genomic-characterisation-of-an-emergent-sars-cov-2-lineage-in-the-uk-defined-by-a-novel-set-of-spike-mutations/563). Among these changes are those that significantly affect the binding process to the host cell: N501Y in the receptor-binding domain (RBD), which increases binding affinity to ACE2 [29] and P681H in the S1-S2 furin cleavage site, which was shown to promote entry into respiratory epithelial cells and transmission in animal models (Figure 1). The South African variant is characterized by eight mutations in the spike protein, including three at critical residues, K417N, E484K, and N501Y, of the RBD [26]. Apparently, these mutations evolved independently in a recently described, Brazilian P.1 (also known as B.1.1.28) variant (https://virological.org/t/genomic-characterisation-of-an-emergent-sars-cov-2-lineage-in-manaus-preliminary-findings/586). Notably, the E484K mutation of these variant has been associated with reduced potency of anti-spike neutralizing antibodies [30]. The joint appearance of multiple spike mutations increases concern that new super-spreading variants are on the rise.

**Figure 1.**
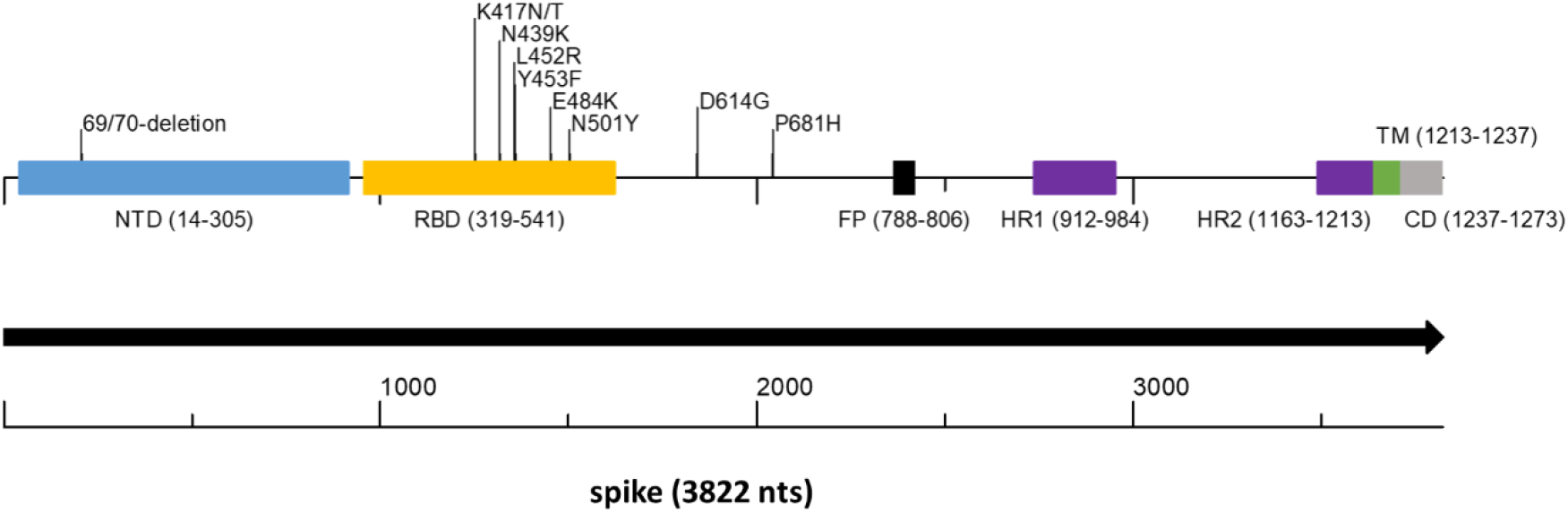
Spike gene scheme with concerning mutations. The spike protein encoded by the S gene is a large protein of 1237 amino acids that contain key structural motives. The figure shows the places of these motives as well as the location of key amino acid changes that occur in important variants. In parenthesis, the start and end of each motive. NTD - N terminal domain (light blue); RBD - Receptor Binding Domain (yellow); FP - Fusion Peptide (black); HR1 and HR2 - Heptapeptide Repeat Sequence 1 and 2 (purple); TM - Trans Membrane (green); CD - Cytoplasmic domain (grey).

The spike protein is the target of SARS-CoV-2 neutralizing antibodies of which over 90 percent target its receptor binding domain (RBD) [31]. Therefore, the SARS-CoV-2 spike glycoprotein has been selected as the primary target of vaccines [32] and anti-viral drugs [10]. Consequently, every new mutation in the spike gene is a potential threat to vaccine and drug efficacy. Clearly, the spike protein’s crucial part in both infectivity and neutralization, and its high tendency to mutate make it the most important sequencing target for monitoring circulating viruses.

Affirming the notion that prompt identification of new variants via genome sequencing is key to outbreak control; emergency public health sequencing initiatives were launched worldwide. The CDC SPHERES (SARS-CoV-2 Sequencing for Public Health Emergency Response, Epidemiology, and Surveillance) program in the US and its ECDC parallel in Europe (European Centre for Disease Prevention and Control. Sequencing of SARS-CoV-2) as well as the COVID-19 Genomics Consortium UK (CoG-UK, https://www.cogconsortium.uk/) are leading examples. A US group has reported monitoring the GISAID (http://www.gisaid.org/) repository database of worldwide SARS-CoV-2 genomic sequences for mutations in the spike gene, associated with frequency shifts at regional and global levels (https://cov.lanl.gov/), [6]

In this work, we present HiSpike, a simple, high-throughput and cost efficient, 3-step targeted NGS method for full sequencing of the spike encoding gene of SARS-CoV-2. HiSpike can be readily adapted to the workflow of standard laboratories engaged in sequencing. We demonstrate that HiSpike provides results per the spike gene identical to results obtained using the protocol for whole genome sequencing of SARS-CoV-2, which was developed by the ARTIC consortium (https://artic.network/ncov-2019). We show that HiSpike reliably detects major clade defining mutations in the spike gene such as the UK 20I/501Y.V1 and the South Africa 20H/501Y.V2 variants. While the ARTIC protocol is an expensive, labor-intensive process, commonly (for Illumina users) requires 4-5 days, the HiSpike method is easily implemented, and obtains sequences in less than 2 days and at a fraction of the cost. Taking into consideration that HiSpike is used for the spike gene only (12.8% of the complete genome), many more samples can be sequenced simultaneously, thus leading to significantly increased capacity.

## RESULTS

With the objective to detect on a national scale the clinical and public health SARS-CoV-2 variants of high concern, we set out to assemble a sequencing protocol with the following guidelines: The protocol must be high throughput, accessible to standard laboratories that engage in sequencing, inexpensive, simple, and with a short turnaround time. Because all current and likely future SARS-CoV-2 variants of high concern are based on spike gene mutations (Figure 1) we focused the sequencing to this region.

To achieve this goal, the HiSpike method for high throughput sequencing of the SARS-CoV-2 spike gene was developed. In this method, nucleic acid (NA) samples were converted to spike libraries for sequencing on a MiSeq (Illumina) instrument, which is among the most common NGS sequencing platforms used worldwide. The HiSpike method can generate sequence data from NA samples within 30 hours and involves three simple steps: two consecutive reactions, RT-PCR1 and PCR2, and a single cleanup step of the pooled MiSeq library (Figures 2A and B). The RT-PCR1 converts the viral spike RNA to cDNA, which is amplified using tailed primers to produce spike amplicons with forward and reverse universal tails (Figure 2B). The spike annealing sites of these primers were primarily derived from the established ARTIC V3 primers for tiling amplicons along the SARS-CoV-2 genome. Likewise, the RT-PCR1 was conducted in two multiplexed reactions to produce two overlapping sets of ∼400 bp spike gene amplicons enclosed by forward and reverse universal tails. These amplicons were combined and subjected to PCR2, which added unique dual indexes and the Illumina P5 and P7 flow cell adaptors to each sample (Figure 2B).

**Figure 2.**
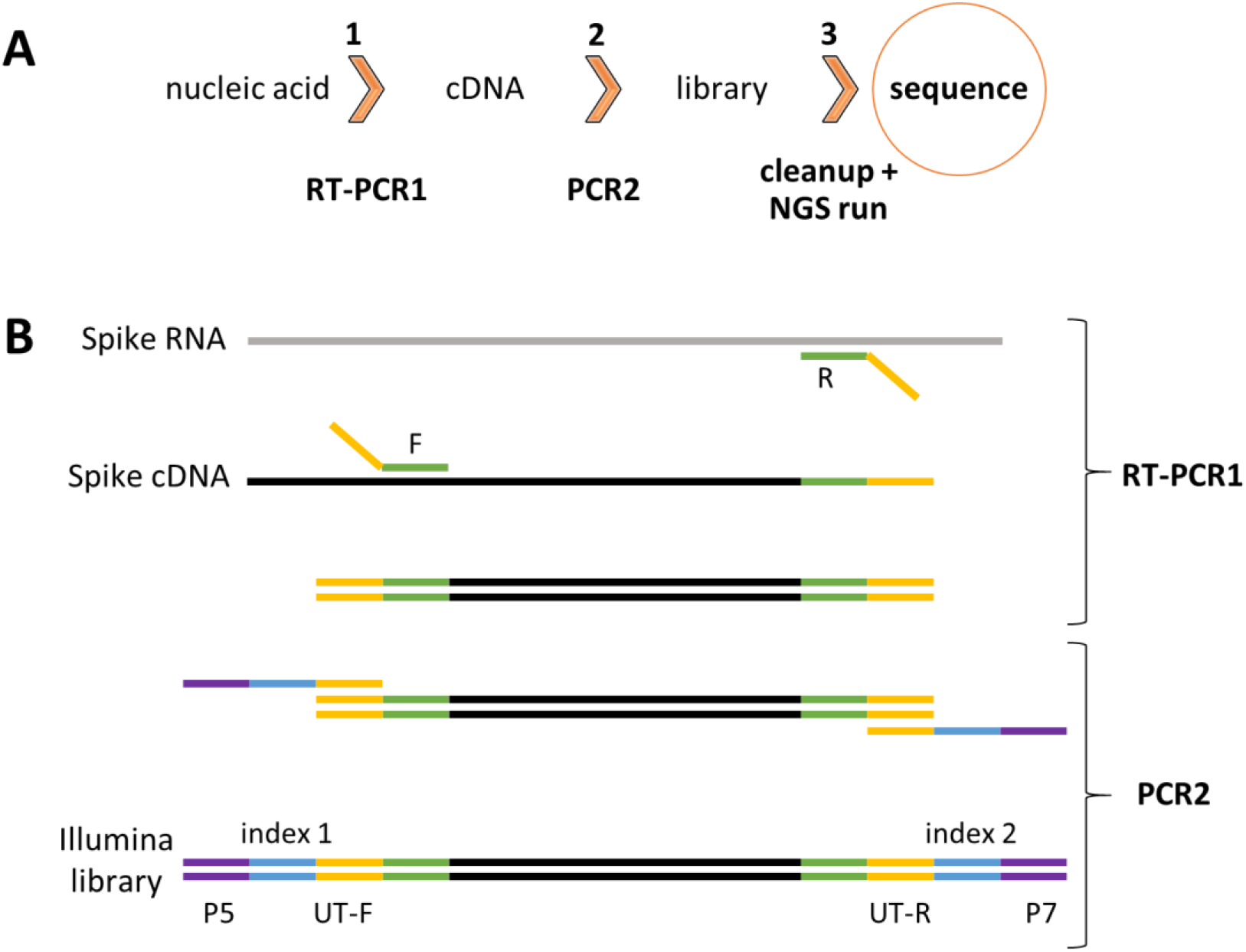
HiSpike method illustration. A. HiSpike three-step protocol outline, B. HiSpike RT-PCR1 and PCR2 Illumina library preparation.

A series of experiments were initially done to optimize and simplify reaction conditions, primer properties, and enzymatic settings to robustly produce high quality libraries of the HiSpike method (data not shown). The optimized RT-PCR1 conditions eliminated the need for equilibration between samples due to the limited primer levels (0.045 µM) that were maxed out during the 45 PCR cycles. Consequently, similar dsDNA amplicon levels in each sample were found and roughly maintained throughout the sequencing process (An example of a MiSeq sample read distribution is shown in Supplementary file S1). In this study, all HiSpike libraries were sequenced with MiSeq’s smallest and fastest flow cell the V2 Nano kit. In a typical 250-bp paired-end run, we obtained about 2 million reads and could simultaneously sequence 96 samples.

To assess the sequencing performance of the HiSpike method, we compared its sequence results from 90 positive SARS-CoV-2 samples with those obtained using the ARTIC V3 protocol on the same samples. These samples spanned a variety of viral loads indicated by their Ct values. The spike sequences generated by the ARTIC method were compared with those of the HiSpike method.

Notably, the spike sequences generated by HiSpike represented various strains (Figure 3 and Supplementary file S6) that differentiated into the similar Nextclade branches as those assigned via ARTIC V3 whole genome sequencing (data not shown). Among the 90 samples 88 showed identical sequences (Figure 3 displaying comparisons of the samples with high sequence coverage) and only 2 samples showed a single nucleotide difference. These differences appeared in samples with high Cts (28 and 30), indicating low levels of viral NA in the original samples. Taken together, these results demonstrate high confidence in the sequence quality obtained by HiSpike.

**Figure 3.**
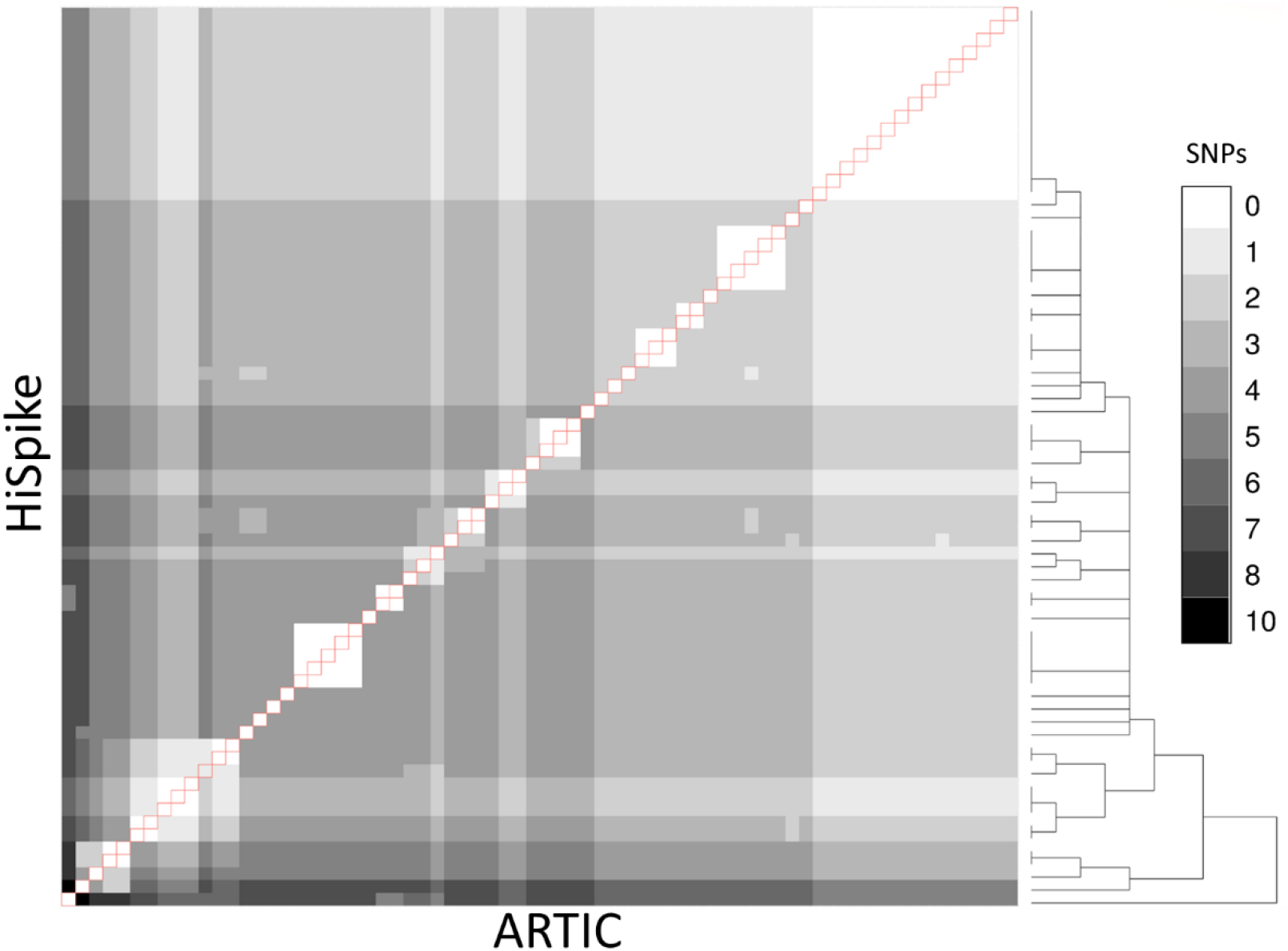
Spike gene sequence comparison between ARTIC and HiSpike. Sequences of 70 samples (with > 90% spike sequence coverage in both ARTIC and HiSpike methods) were compared using a heat-map. Grey shades between white and black indicate 0 to 10 SNPs respectively. HiSpike sequences were aligned based on an hierarchical clustering tree (shown to the right) vertically and the ARTIC sequences of the same samples were aligned horizontally. Pairs of the same sample are represented in the diagonal line (outlined in red).

The validated HiSpike sequencing capacities and short turnaround time from NA to sequence led to the selection of this method for nationally urgent sequence assignments of positive COVID-19 cases. These included travelers arriving from countries with a high incidence of spike variants of concern, instances of reinfection, severely ill patients, and hospitalized pregnant women. A total of 304 clinical NA samples arrived on five different occasions during two critical weeks in January (Jan. 14 to 28, 2021). During this time, a global spread of high concern variants (e.g. UK, South African, and Brazilian P.1) forced lockdowns in many countries including Israel, while massive COVID-19 immunization efforts took place. Samples were processed upon arrival and the average time from sample to sequence was less than 30 hours. The HiSpike method identified 119 UK variants and 7 South African variants among other multiple clade branches (Figure 4 and Supplementary file S6). These findings demonstrate that HiSpike can sequence essentially any spike variant in a near real-time manner.

**Figure 4.**
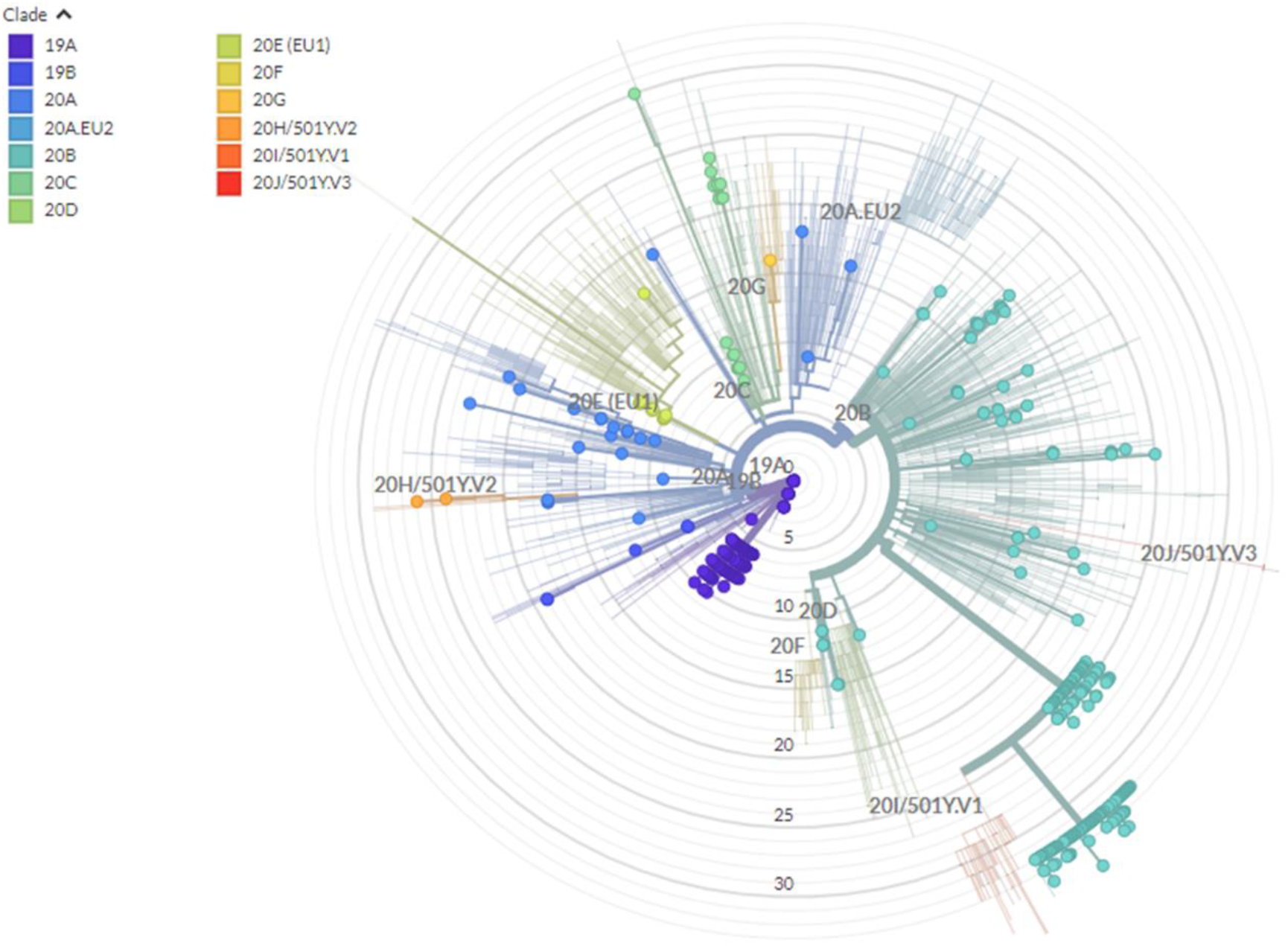
Phylogenetic tree based on spike gene sequences generated by HiSpike method. Spike gene sequences of 396 samples generated by HiSpike method were uploaded to Nextclade platform. SARS-CoV-2 clades are illustrated by different colors and locations on the radial tree. HiSpike sequences are represented by the full circles on a backgound of the Nextcalde’s global clade tree.

To determine HiSpike’s limit of detection, we plotted the Ct value (as the viral load indicator) and spike gene sequence coverage breadth. To this end, 394 samples (comprised of the validation sample set and the urgent clinical samples) with Ct values ranging from 11 to 40 were analyzed. The median coverage of samples were as follows: Ct 11 - 25, Ct 25 – 30, Ct 30 – 35, and Ct 35 -40 were 99.3%; 93.0%; 47.3%; and 17.3% respectively (Figure 5A). Notably, samples in this study were collected form several clinical laboratories that use various extraction systems and SARS-CoV-2 determination methods. In rare cases, we observed low coverage in samples with Cts below 25 (Figure 5A) which likely represent poor quality or degraded NA that in our experience are also apparent when using the ARTIC method.

**Figure 5.**
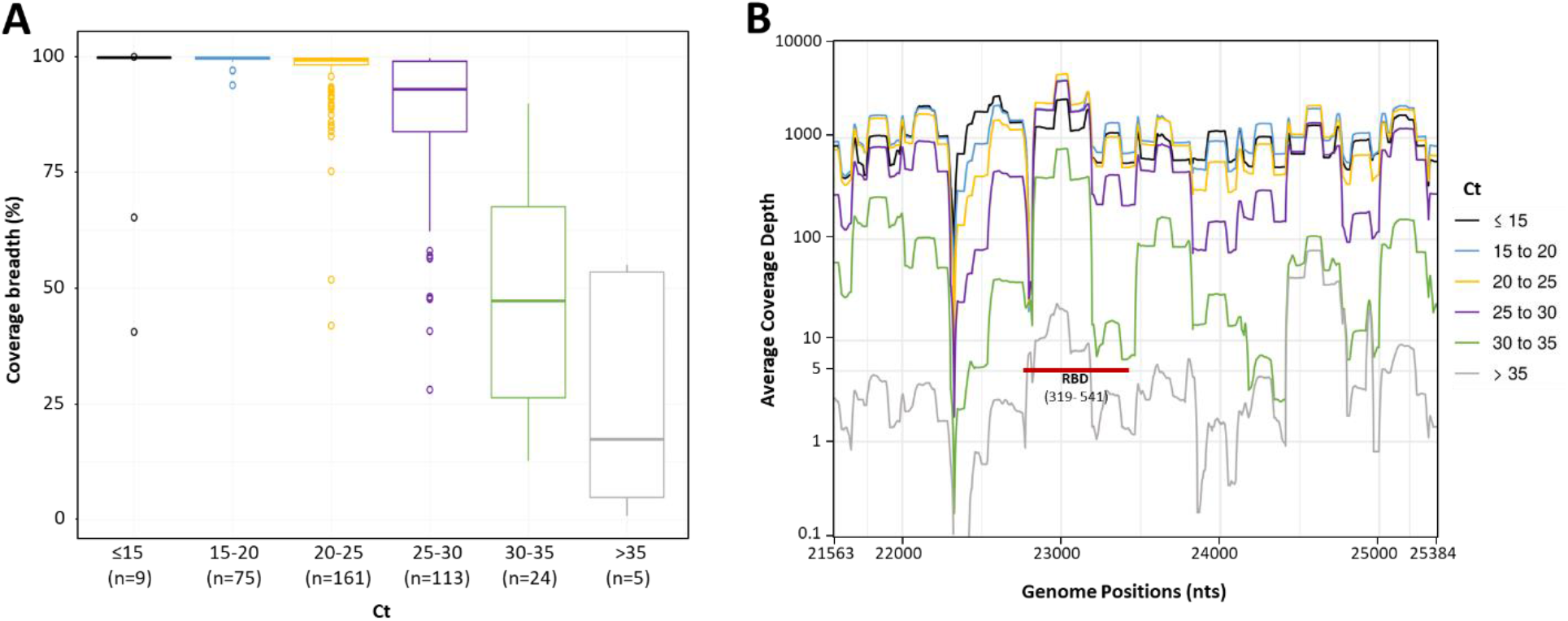
Spike sequence breadth and depth as a function of different sample Ct levels. Spike gene sequences of 396 samples were divided to 6 groups based on their Ct value (up to 15, 15-20, 20-25, 25-30, 30-35, and over 35). The number (n) of sample in each group is in brackets. A. The percentage of spike gene coverage, with at least 5 reads for different Ct levels is shown by Boxplots. The Boxplots represent the coverage breadth value distributions from the lower to the upper quartiles. The inner horizontal line indicates the median. The vertical whiskers extend to the most extreme data point which is no more than 1.5 times the interquartile range. More extreme data points are drawn as circles. B. Sequence coverage depth along the spike gene. The average coverage depth was plotted, on a logarithmic scale, along the genome positions. Lines were smoothed by a sliding window of length 20. At coverage depth x5 the spike gene region encoding the receptor binding domain is marked with a red line and annotated (RBD and in brackets its amino acid location).

We next assessed the average coverage depth along the spike gene with relation to its Ct. This parameter is important because it provides insights into the coverage expectancy along the spike sequence. As seen in Figure 5B, the average coverage depth along the spike gene varied. While at Cts of 25 – 30, the average coverage depth was more than 100 at Cts above 30, there was a significant decline and more regions remained below the five independent reads cut off (Figure 5B). Notably, key parts of the RBD in the spike gene were amplified and sequenced at coverage of 5 reads or more even in samples with high Cts (over 30), allowing detection of mutations like N501Y (Figure 5B, RBD red bar).

We conclude that the HiSpike method is highly robust and suitable for full spike gene sequencing of samples with Cts up to 30. This is remarkably similar to the specifications of all other ARTIC based sequencing methods for SARS-CoV-2 [33]. Importantly, key regions in the spike gene will likely be detected also in samples with very low virus content.

## MATERIALS AND METHODS

For user convenience, we included an instruction manual for implementing the HiSpike method in Supplementary document S2.

### Sample source and characterization

Positive COVID-19 samples of either primary nasopharyngeal swabs or extracted NA were collected from various COVID-19 laboratories in Israel. NA samples were stored at −80°C prior to use and primary samples were kept at 4°C (up to 72 hours) prior to extraction. Viral inactivation of swab samples was performed using Lysis Buffer (Seegene) at a volume ratio of 0.75 (buffer : sample) for 10 minutes at room temperature. Total NA samples were extracted using automated instruments of MagNA Pure (Roche) or magLEAD (Precision System Science Co., Ltd.).

The cycle thresholds (Cts) of SARS-CoV-2 positive samples were determined either by the source lab or, in unknown cases, in house using Allplex™ 2019-nCoV qPCR kit (Seegene). The qPCR reactions were performed according to manufacturer’s instructions. Briefly, reactions were comprised of 25 µL, consisting of 17 µL master mixes (Buffer, RNase-free water and 2019-nCoV MOM and Enzyme) and 8 µL NA templates. Reactions were added to 96-well plates each of which included a positive and negative control. The qPCRs were performed on a CFX96 Touch™ instrument (Bio-Rad) with the following conditions: 20 minute reverse transcriptase step at 50°C, 15 minute heat-start step at 95°C, and 45 cycles of 15 second denaturing period at 95°C followed by a combined annealing and extension step at 58°C for 30 seconds. Finally, the Ct values of E, RdRP, and N genes were determined using the Seegene Viewer software (Seegene). For simplicity, in this work we used the average Ct of all three genes.

### HiSpike Library preparation

Primers used in this study were manufactured by metabion or IDT and are shown in Supplementary files S3 and S4.

RT-PCR1 primers contained 3’ annealing sequences corresponding to sites along the spike gene and 5’ forward or reverse universal tails (denoted in the sequence below by upper case letters). A primer binding site, an Illumina component for initiation of reads 1 and 2 (denoted below with lower case letters), was located between the universal tails and the annealing regions resulting in the following strands: 5’TCGTCGGCAGCGTCagatgtgtataagagacag[spike annealing sense sequence]3’and 5’GTCTCGTGGGCTCGGagatgtgtataagagacag[spike annealing antisense sequence] 3’ for the forward and reverse primers respectively (Supplementary file S3). The spike annealing sites (indicated in square brackets) were mostly derived from the SARS-CoV-2 V3 ARTIC primers for tiling amplicons (https://artic.network/resources/ncov/ncov-amplicon-v3.pdf) and in some cases, additional primers with alternative annealing sequences were added to increase the sequencing coverage (Supplementary file S3).

The PCR2 primers termed F-P5 (i01 – i12) and R-P7 (i01 – i08) contained the following sequences: 5’AATGATACGGCGACCACCGAGATCTACAC[i5]tcgtcggcagcgtc3’ and 5’CAAGCAGAAGACGGCATACGAGAT[i7]gtctcgtgggctcgg3’ respectively (Supplementary file S4). Each primer in the primer pair harbored a unique 8 base index, denoted i5 or i7, and a 3’ sequence (in lower case letters) of the forward and reverse universal tails that anneal and amplify the RT-PCR1 products, thereby adding the dual index and unidirectional P5 and P7 termini.

#### Step 1 of the HiSpike method - library preparation: RT-PCR1

Two 10 µL multiplex RT-PCR1 reactions were composed of either primer mix 1 or mix 2 using SensiFast (Meridian Bioscience). The reaction consisted of 5 µL 2x SensiFAST, 0.4 µL RiboSafe RNase Inhibitor, 0.2 µL Reverse transcriptase, 1.7 µL H_2_O, and 1 µL of primer mix set 1 or 2 (0.45 µM of each primer), and 3 µL NA template. The RT-PCR was performed on a Biometra TOne 96 Standard Thermal Cycler instrument (Analytik Jena) with the following conditions: 10 minute reverse transcriptase step at 45°C, 4 minute heat-start step at 95°C, and 45 cycles of 15 seconds denaturing period at 95°C, annealing of 30 seconds at 60°C and elongation at 72°C for 30 seconds. The products of corresponding samples from reaction sets 1 and 2 were combined and used as the template for PCR2.

#### Step 2 of the HiSpike method - library preparation: PCR2

PCR2 was performed using KOD Hot Start DNA Polymerase (Merck Millipore). Reactions were set to 15 µL and contained 4 µL of each of the uniquely indexed primers F-P5 (1.2 µM) and R-P7 (1.2 µM), 1.0 µL RT-PCR template and the reaction mix of 1.5 µL 10X Buffer, dNTPs (2 mM each) and MgSO_4_ (25 mM), 1.2 µL H_2_O, and 0.3 µL KOD Hot Start DNA Polymerase (1 U/ µL). Reactions were performed on a Biometra TOne 96 Standard Thermal Cycler instrument (Analytik Jena) with the following conditions: 4 minutes hot-start step at 95°C, and 15 cycles of 15 seconds denaturing period at 95°C, annealing of 30 seconds at 58°C and elongation at 72°C for 30 seconds.

### Library cleanup and MiSeq loading (Step 3 of the HiSpike method)

Products of PCR2 were pooled by collecting 4 µL from each well. The pooled library was diluted 1 : 3 (sample : H_2_O) and purified using the ProNex® Size-Selective Purification System (Promega) at a ratio of 1.4 : 1 (ProNex® chemistry : sample) according to the manufacturers’ instructions. In Short, 70 µL beads and 50 µL diluted sample were mixed and incubated for 10 min and placed on a magnetic stand for 2 minutes. The supernatant was discarded and the resin was washed twice by two consecutive 1-minute washes using 200μl of Wash Buffer each. After discarding the second portion of wash buffer, the resin was allowed to air-dry for 5 minutes. It was then removed from the magnetic stand and resuspended with 50 µL elution buffer for 5 minutes. Finally, the sample was placed on the magnetic stand for 1 minute and the eluted purified HiSpike library was collected. The dsDNA concentration of the purified HiSpike library was determined by Denovix QFX Fluorometer, using the DeNovix dsDNA High Sensitivity assay.

The HiSpike library, with an expected fragment average size of 550 bp, was examined by agarose gel electrophoresis (1.7%) and on the Fragment Analyzer 5200 (Agilent) with HS NGS Fragment (1-6,000bp) kit. The purified library was diluted to 4 nM and denatured by mixing 5 µl of library with 5 µL 0.2 N NaOH for 5 minutes. The denatured library was further diluted to 12 pM and a 1% PhiX control (PhiX Control v3) was added. Sequencing of all HiSpike samples in this study were performed on an Illumina MiSeq platform using a 250-bp paired-end read v2 Nano kit (Cat. # MS-103-1003). For user convenience, we included a typical MiSeq sample sheet for 96 indexes in Supplementary file S5.

### ARTIC V3 protocol for SARS-CoV-2 full genome sequencing

RNA in extracted NAs was reverse transcribed to single strand cDNA using SuperScript IV (ThermoFisher Scientific, Waltham, MA, USA) as per manufacturer’s instructions. SARS-CoV-2 specific primers designed to capture SARS-CoV-2 whole genome (version 3, https://github.com/artic-network/artic-ncov2019/blob/master/primer_schemes/nCoV-2019/V3/nCoV-2019.tsv) - total 218 primers, divided into two primer pools designed by Josh Quick from ARTIC Network) were used to generate double strand cDNA and amplify it via PCR using Q5 Hot Start DNA Polymerase (NEB) [34]. Briefly, each sample underwent two PCR reactions with primer pool 1 or 2 and 5X Q5 reaction buffer, 19 mM dNTPs and nuclease-free water. The resulting DNA was combined and quantified with a Denovix QFX Fluorometer, using the DeNovix dsDNA High Sensitivity assay as per manufacturer’s instructions, and 1ng of amplicon DNA in 5 µL per sample was used for the library preparation. Libraries were prepared using the NexteraXT library preparation kit and NexteraXT index kit V2 as per manufacturer’s instructions (Illumina, San Diego, CA, USA). Pooled libraries were purified with AMPure XP magnetic beads (Beckman Coulter, Brea, CA, USA) and the pooled library concentration was determined by Denovix QFX Fluorometer, using the DeNovix dsDNA High Sensitivity assay. The pooled library validation and mean fragment size was determined by Fragment Analyzer 5200 (Agilent) with the HS NGS Fragment (1-6,000bp) kit. The mean fragment size was ∼350 bp, as expected. The molarity concentration of the library was calculated and diluted to 4 nM, denatured and further diluted to 12 pM. Finally, the sample was loaded on MiSeq Reagent Kit v3 (600-cycle, Cat. # MS-102-3003) and a paired end 151X2 program was set.

### Sequence clean-up, mapping and determination of coverage (Bioinformatics analysis)

Trimmomatic-0.36 was used for quality trimming of the raw read sequences, by clipping 3’ read ends with sliding window quality lower than 15 in a window of 4 nucleotides, and at the same time trimming the 5’ primer sequence used for targeting the virus genome, by cropping 30 nucleotides from the head of the read. Reads with less than 50 bases remaining length were filtered out. The resulting reads were mapped to the spike gene of the Wuhan-Hu-1 reference genome (NCBI Reference Sequence NC_045512.2) using bwa v0.7.12-r1039 MEM algorithm. Samtools 1.10 was used for sorting, merging pileup, and consensus sequence was created with ivar 1.0 with minimum quality score threshold to count base of 15, and minimum depth to call consensus of 5. Point coverage depth was determined with samtools depth with base quality threshold of 15 and mapping quality threshold of 15. Coverage breadth was defined as the ratio of the reference sequence covered by X5 depth or more. FASTA sequences were uploaded to Nextclade (https://clades.nextstrain.org/), which allowed various options for visualization of sequence relationship to other sequences in the batch, or to other sequences representing the currently known clades. For each sequence, Nextclade provided a complete list of established and new spike gene mutations, both in nucleotide and amino acid notation.

## DISCUSSION

The SARS-CoV-2 virus will surely continue to evolve over time in human populations. Real-time monitoring of the circulating strains by sequencing is essential to combat the current pandemic. The spike protein centrality in viral infectivity, transmissibility, and reactivity with neutralizing antibodies is the current most important element for sequence monitoring. Globally, ARTIC’s method for full genome sequencing of SARS-CoV-2 has become predominant. While serving as an essential tool for genomic epidemiology, this method is both labor intensive, and expensive and consequently it is unfit for the current real time high-load sequencing demands. The HiSpike method, which follows mutations in the spike gene only, is more suitable for such tasks. Using this method, one can produce high quality sequencing libraries in three steps, RT-PCR1, PCR2, and NA pool cleanup making the assay simple and inexpensive.

The RT-PCR1 primer annealing sites along the spike gene were derived from the well-established ARTIC primers (https://www.protocols.io/view/ncov-2019-sequencing-protocol-v3-locost-bh42j8ye), plus newly designed primers to achieve adequate coverage. The 5’ end of these primers contained forward and reverse universal tails. The tail design of RT-PCR1 and PCR2 primers shares similar characteristics with our patent pending method (Nissan, I, el al., 2018. MOLECULAR TYPING OF MICROBES. USA patent 62715813). Notably, Gohl et al [33] also incorporated these adapter tails to ARTIC V3 primers for full genome sequencing. Although, Gohl’s method required four PCR pools instead of the original ARTIC two pools, it significantly reduced the cost and labor of classic ARTIC methods.

Here we show that HiSpike is a much simpler method than any currently published method, that: 1) requires (similar to original ARTIC version) only two pools per sample, 2) generates cDNA and target amplification in a single RT-PCR1 step, and 3) does not require library normalization because it is achieved by substantial consumption of the limited levels of PCR1 primers. HiSpike is also flexible, as it requires reagents that are commonly available from various manufactures. In this study, we used SensiFAST™ SYBR Hi-ROX Kit (Cat. # BIO-92005) and KOD Hot Start DNA Polymerase (Cat. # 71086) for RT-PCR1 and PCR2 respectively. Yet, high quality libraries were also obtained with Xpert One-Step RT-PCR Kit (grisp Cat. GE50.0100) for RT-PCR1 and TaKaRa Ex Taq (TaKaRa Cat. # RR001A) for PCR2 (data not shown). The simplicity and reagent flexibility together with its low cost make HiSpike significantly more accessible and suitable for a wider community.

HiSpike is also remarkably useful for high-throughput sequencing. It can sequence 96 samples using the cheapest and fastest MiSeq’s Nano kit. In a typical run, we obtained about 2 million reads. The MiSeq Reagent Kit V2 (500-cycles) (Cat. # MS-102-2003) offers more than 10 fold reads per run (24-30 million), therefore, at the same cluster density, using this kit, it is possible to sequence nearly 1000 samples per flow cell. Other whole genome sequencing methods ([33], and ARTIC V3) require the most expensive MiSeq Reagent Kit V3 (600-cycle) (Cat. # MS-102-3003) that gives 44–50 million reads per run. Using HiSpike we can expect to sequence up to 2400 samples in a single MiSeq Reagent Kit V3 run. Overall, we estimate that the HiSpike method reduces cost per sample to about 10% of that using ARTIC.

Recently, spike protein multi-mutation variants have appeared with high transmissibility and potential resistance to neutralization by either convalescent or vaccine-elicited sera. The current record high infection rates coupled with massive vaccination campaigns are expected to give rise to emergence of additional spike variants. Therefore, it is important to have a tool for rapid and high-throughput sequencing. The ARTIC whole genome sequencing methods are effective for high definition epidemiologic purposes in which short time to sequence and high volume is not the primary desire. Although the ARTIC sequencing method when performed using the nanopore technique (ARTIC’s original platform) can achieve sample to sequence in less than 24 hours it is not high-throughput and its high price per sample is prohibitive for many standard laboratories. HiSpike can fill in the need for vast sequencing to cover a national scale of all SARS-CoV-2 positive samples: it is simple to execute, high-throughput and an inexpensive method that produces near real-time results.

Because our method requires less reads per sample, it also saves expensive storage space and computation resources for analysis. Essential web platforms like GISAID (https://www.gisaid.org/) allow submission of complete or nearly complete genomes, while they also perform multiple analyses that are focused on the spike gene such as the spike glycoprotein mutation surveillance. However, a most needed spike dedicated database for sequence deposit and global sharing for comparison and variant alerts has yet to be establish.

We have enumerated the many strengths of HiSpike; its desirable features and benefits that make it simple, rapid, high-throughput, affordable, and a key tool to assist formulating public health policy. Nevertheless, HiSpike does have some limitations, chief among them being that it does not sequence genomic regions outside the spike gene, which are covered by other protocols for whole genome sequencing [33, 35-37]. Rarely, mutations outside the spike gene can influence transmissibility. However, the most important aspect of this limitation is that molecular epidemiology including its uses for clade definition and outbreak profiling to aid contact tracing, is mostly effective at the whole genome level.

To the best of our knowledge, HiSpike is currently the only documented sequencing tool for specific monitoring of spike mutations. To ease transitions to the HiSpike method, we included a detailed user instruction manual (Supplementary file S2). We propose using HiSpike for routine sequencing of positive SARS-CoV-2 samples for near real-time monitoring of emerging spike mutations. In this manner, public health resources can help contain potential super spreading or vaccine-escape variants, even before these variants cause frequency shifts at the local, regional, or global levels.

## Data Availability

The authors confirm that the data supporting the findings of this study are available within the article and its supplementary materials.

## Authors contributions

F.E., conceived the HiSpike concept, conceived and designed the experiments, conducted experiments, analyzed data, and wrote the manuscript;

V.Z.G., conceived and designed the experiments, conducted experiments and helped write the manuscript;

R.M., analyzed data and helped write the manuscript;

F.P.J., wrote much of, and critically reviewed, the manuscript;

R.S., Critical reading and writing of the manuscript;

K.I., W.R. and B.T.N. conducted the experiments and helped write the manuscript;

D.Z., Critical reading of the manuscript;

B.E., Conducted experiments and analyzed data

Z.N., Analyzed data and critical reading of the manuscript

M.O., Analyzed data and critical reading of the manuscript

M.E., Project supervision and critical reading

R.E., Project supervision, conceived and designed the experiments, and critical reading of the manuscript;

N.I., conceived the HiSpike concept, conceived and designed the experiments, conducted experiments, analyzed data, and wrote the manuscript;

All authors read and approved the final manuscript.

## SUPPLEMENTARY FILE LIST

**Supplementary file S1**. Percentage of clusters passing filter (PF) for 92 samples of a MiSeq (Illumina) run. The Red column represents the average PF reads/sample.

**Supplementary file S2**. A user manual for the HiSpike method for sequencing 96 samples of the SARS-CoV-2 spike gene.

**Supplementary file S3**. List of HiSpike RT-PCR1 primer sequence and composition.

**Supplementary file S4**. List of indexing PCR primer (PCR2) sequences and composition used in this study for HiSpike method.

**Supplementary file S5**. Example of a SampleSheet for MiSeq run in a tab delimited (cs) format.

**Supplementary file S6**. Nextclade (https://clades.nextstrain.org) analysis results of 396 samples that were sequenced by the HiSpike method. Sample IDs S-0001 -S0090 were (collected before December, 2020) were sequenced by HiSpike and ARTIC methods for validation. Sample IDs S-0091 - S-0396 were sequenced by HiSpike as urgent field samples (between 14 – 28 January of 2021). Samples with UK (20I/501Y.V1) signatures are marked in green and those of the South African (20H/501Y.V2) in brown.

