## Supplementary file S2 for "HiSpike: A high-throughput cost effective sequencing method for the SARS-CoV-2 spike gene"

[1]National Public Health Laboratory Tel Aviv, Israel Ministry of Health, Israel

[2]Central Virology Laboratory, Israel Ministry of Health, Chaim Sheba Medical Center, Tel-Hashomer, Ramat Gan, Israel

### HiSpike Method for Sequencing 96 SARS-CoV-2 positive samples

*Reagents and Consumables*

1. HiSpike Primers - sequences are detailed in Supplementary file S3
2. Index primers - sequences are found in Supplementary file S4
3. Prepare a 4.5 µM (x100) stock of HiSpike Primers for primer pool-1 and primer pool-2 according to the tables below:
   **Note**: the primer mix stocks volume of 445 µL (each) will be enough for sequencing of ~ 400 samples and should be stored between uses at -20˚ C.
4. SensiFast One-Step Kit
5. KOD Hot Start DNA Polymerase Kit
6. PCR-grade water
7. 96-well PCR plates
8. Microseal adhesive seals
9. ProNex® Size-Selective Purification System
10. Magnetic separation plate.

*HiSpike primer mix preparation of 4.5 µM (x100 stock)*

Prepare HiSpike primer pool-1 and primer pool-2 according to the tables below:
(The HiSpike primer sequences can be found in Supplementary file S3)

| **Primer Pool 1** | |
|---|---|
| **4.5 µM (each)** | |
| **primer ID (100 µM)** | **(µL)** |
| 1_F_HiSpike | 20 |
| 1_R_HiSpike | 20 |
| 3_F1_HiSpike | 20 |
| 3_F2_HiSpike | 20 |
| 3_R1_HiSpike | 20 |
| 3_R2_HiSpike | 20 |
| 5_F1_HiSpike | 20 |
| 5_F2_HiSpike | 20 |
| 5_R1_HiSpike | 20 |
| 5_R2_HiSpike | 20 |
| 7_F_HiSpike | 20 |
| 7_R_HiSpike | 20 |
| 9_F_HiSpike | 20 |
| 9_R_HiSpike | 20 |
| 11_F_HiSpike | 20 |
| 11_R_HiSpike | 20 |
| 13_F1_HiSpike | 20 |
| 13_F2_HiSpike | 20 |
| 13_R1_HiSpike | 20 |
| 13_R2_HiSpike | 20 |
| 15_F_HiSpike | 20 |
| 15_R_HiSpike | 20 |
| DDW | 5 |
| **total volume** | 445 |
| **sum µM (each)** | 4.5 |

| **Primer Pool 2** | |
|---|---|
| **4.5 µM (each)** | |
| **primer ID (100 µM)** | **(µL)** |
| 2_F_HiSpike | 20 |
| 2_R_HiSpike | 20 |
| 4_F_HiSpike | 20 |
| 4_R_HiSpike | 20 |
| 6_F1_HiSpike | 20 |
| 6_F2_HiSpike | 20 |
| 6_R1_HiSpike | 20 |
| 6_R2_HiSpike | 20 |
| 8_F1_HiSpike | 20 |
| 8_F2_HiSpike | 20 |
| 8_R1_HiSpike | 20 |
| 8_R2_HiSpike | 20 |
| 10_F_HiSpike | 20 |
| 10_R_HiSpike | 20 |
| 12_F_HiSpike | 20 |
| 12_R_HiSpike | 20 |
| 14_F_HiSpike | 20 |
| 14_R_HiSpike | 20 |
| 16_F_HiSpike | 20 |
| 16_R_HiSpike | 20 |
| DDW | 45 |
| **total volume** | 445 |
| **sum µM (each)** | 4.5 |

Note: the primer mix stocks volume of 445 µL (each) will be enough for sequencing of ~ 400 samples and should be stored between uses at -20˚ C.

*HiSpike protocol for 96 wells*

### Step 1: HiSpike library preparation: RT-PCR1

1. Prepare the following RT-PCR1 reaction mix (RM1) and keep on ice:

| RT-PCR1 reaction mIx (RM1) for 10 µL reaction | | |
|---|---|---|
| | reaction Pool x | |
| **reagent** (SensiFast One-Step) | **1** | **210** |
| 2x sensiFAST (µL) | 5 | 1050 |
| reverse tracscriptase (µL) | 0.1 | 21 |
| Ribosafe RNAse inibitor (µL) | 0.2 | 42 |
| DDW (µL) | 1.7 | 357 |
| sum | 7 | 1470 |

Note: the pool of 210 reactions is calculated for 96 samples x 2 plates (=192) with an addition of 10%

2. Divide RM1 into two pools each containing 724 µL
3. Add to each pool 10.5 µL of either PM-1 (4.5 µM) or PM-2 (4.5 µM), mix and keep on ice.
4. Prepare two 96 PCR plates, one for each pool, and add to each well 7 µL of either pool with the added PM-1 or PM-2. Keep PCR plates on ice.

| | PM-1 | | | | | | | | | | | |
|---|---|---|---|---|---|---|---|---|---|---|---|---|
| | **1** | **2** | **3** | **4** | **5** | **6** | **7** | **8** | **9** | **10** | **11** | **12** |
| **A** | 1 | 9 | 17 | 25 | 33 | 41 | 49 | 57 | 65 | 73 | 81 | 89 |
| **B** | 2 | 10 | 18 | 26 | 34 | 42 | 50 | 58 | 66 | 74 | 82 | 90 |
| **C** | 3 | 11 | 19 | 27 | 35 | 43 | 51 | 59 | 67 | 75 | 83 | 91 |
| **D** | 4 | 12 | 20 | 28 | 36 | 44 | 52 | 60 | 68 | 76 | 84 | 92 |
| **E** | 5 | 13 | 21 | 29 | 37 | 45 | 53 | 61 | 69 | 77 | 85 | 93 |
| **F** | 6 | 14 | 22 | 30 | 38 | 46 | 54 | 62 | 70 | 78 | 86 | 94 |
| **G** | 7 | 15 | 23 | 31 | 39 | 47 | 55 | 63 | 71 | 79 | 87 | 95 |
| **H** | 8 | 16 | 24 | 32 | 40 | 48 | 56 | 64 | 72 | 80 | 88 | 96 |

| | PM-2 | | | | | | | | | | | |
|---|---|---|---|---|---|---|---|---|---|---|---|---|
| | **1** | **2** | **3** | **4** | **5** | **6** | **7** | **8** | **9** | **10** | **11** | **12** |
| **A** | 1 | 9 | 17 | 25 | 33 | 41 | 49 | 57 | 65 | 73 | 81 | 89 |
| **B** | 2 | 10 | 18 | 26 | 34 | 42 | 50 | 58 | 66 | 74 | 82 | 90 |
| **C** | 3 | 11 | 19 | 27 | 35 | 43 | 51 | 59 | 67 | 75 | 83 | 91 |
| **D** | 4 | 12 | 20 | 28 | 36 | 44 | 52 | 60 | 68 | 76 | 84 | 92 |
| **E** | 5 | 13 | 21 | 29 | 37 | 45 | 53 | 61 | 69 | 77 | 85 | 93 |
| **F** | 6 | 14 | 22 | 30 | 38 | 46 | 54 | 62 | 70 | 78 | 86 | 94 |
| **G** | 7 | 15 | 23 | 31 | 39 | 47 | 55 | 63 | 71 | 79 | 87 | 95 |
| **H** | 8 | 16 | 24 | 32 | 40 | 48 | 56 | 64 | 72 | 80 | 88 | 96 |

5. Prepare a sample sheet recording the position of the sample on each plate. Add 3 µL of nucleic acid sample to the appropriate wells on plate of PM-1 and PM-2 (for 10 uL total per well) and seal the cover wells (with caps or plate sealer).

6. Place each plate in a thermocycler and perform the following PCR program:

| **RT-PCR1 program (SensiFast)** | | |
|---|---|---|
| **step** | **$^{o}$C** | **time** |
| 1 | 45 | 10 min |
| 2 | 95 | 2 min |
| 3 | 95 | 15 sec |
| 4 | **60** | 30 sec |
| 5 | 72 | 30 sec |
| 6 | go to 3 x 44 | |

7. Centrifuge Plate-1 and Plate-2 of RT-PCR1 and combine corresponding samples into one plate; use a 10 µL multichannel pipet to pipet all fluid (max 10µL) from plate 1 to plate 2. Note: necessary steps should be implemented to avoid amplicon contamination. We recommend handling in a secluded area.

### Step 2: HiSpike library preparation: PCR2

1. Prepare a PCR 96 well plate for PCR2 and add index primers as follows:

(equences of index primers can be found in Supplementary file S4)

4 µL of index primers F-P5 i01 - i12 (1.2 µM) to each well in columns 1 – 12 respectively
4 µL of index primers R-P7 i01 - i08 (1.2 µM) to each well in rows **A – H** respectively. (For example F-P5 i01 to wells of column 1 and R-P7 i01 to wells of row A as indicated below). To avoid index contamination between columns use new tips for each lane.

| **PCR2** | | | F-P5 i01 | F-P5 i02 | F-P5 i03 | F-P5 i04 | F-P5 i05 | F-P5 i06 | F-P5 i07 | F-P5 i08 | F-P5 i09 | F-P5 i10 | F-P5 i11 | F-P5 i12 |
|---|---|---|---|---|---|---|---|---|---|---|---|---|---|---|
| | | | ↓ | ↓ | ↓ | ↓ | ↓ | ↓ | ↓ | ↓ | ↓ | ↓ | ↓ | ↓ |
| | | | **1** | **2** | **3** | **4** | **5** | **6** | **7** | **8** | **9** | **10** | **11** | **12** |
| R-P7 i01 | → | **A** | 1 | 9 | 17 | 25 | 33 | 41 | 49 | 57 | 65 | 73 | 81 | 89 |
| R-P7 i02 | → | **B** | 2 | 10 | 18 | 26 | 34 | 42 | 50 | 58 | 66 | 74 | 82 | 90 |
| R-P7 i03 | → | **C** | 3 | 11 | 19 | 27 | 35 | 43 | 51 | 59 | 67 | 75 | 83 | 91 |
| R-P7 i04 | → | **D** | 4 | 12 | 20 | 28 | 36 | 44 | 52 | 60 | 68 | 76 | 84 | 92 |
| R-P7 i05 | → | **E** | 5 | 13 | 21 | 29 | 37 | 45 | 53 | 61 | 69 | 77 | 85 | 93 |
| R-P7 i06 | → | **F** | 6 | 14 | 22 | 30 | 38 | 46 | 54 | 62 | 70 | 78 | 86 | 94 |
| R-P7 i07 | → | **G** | 7 | 15 | 23 | 31 | 39 | 47 | 55 | 63 | 71 | 79 | 87 | 95 |
| R-P7 i08 | → | **H** | 8 | 16 | 24 | 32 | 40 | 48 | 56 | 64 | 72 | 80 | 88 | 96 |

Note: there should now be 8 uL per well.

2. Prepare PCR2 KOD Hot Start reaction mix (RM2) for 96 wells as follows and keep on ice:

| **PCR2 reaction mix (RM2) for 15 µL reaction** | | |
|---|---|---|
| | **pool x (µL)** | |
| **reagents** (KOD Hot Start) | **1** | **105** |
| 10x buffer | 1.5 | 158 |
| 25mM Mg | 1.5 | 158 |
| dNTPs (2 mM each), final 0.2 mM each | 1.5 | 158 |
| KOD Hot Start | 0.3 | 32 |
| H2O Sigma W4502 autoclaved | 1.2 | 126 |
| sum | 6 | 630 |

Note: the pool of 105 reactions is calculated for 96 samples with an addition of 10%.

3. Add to each well of the PCR2 plate 6 µL of KOD Hot Start RM and 1 µL template samples of the joint RT-PCR1 to its corresponding well.
   Note: there should now be 15 uL per well.
4. Perform the following PCR2 program:

| **PCR2 program (KOD)** | | |
|---|---|---|
| **step** | **° C** | **time** |
| 1 | 95 | 4 min |
| 2 | 94 | 30 sec |
| 3 | 58 | 30 sec |
| 4 | 72 | 45 sec |
| 5 | go to 2 x14 | |

5. As a check point, it is highly recommended to examine some of the PCR products on an agarose gel (1.7 %) before running the MiSeq sequencing. The expected band sizes for RT-PCR1 and PCR2 are ~450 bp and ~550 bp respectively (Figure 1).

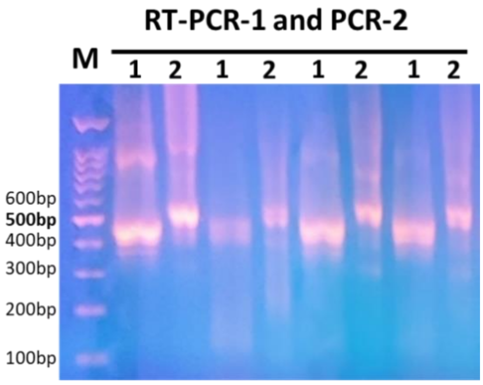


Figure 1. Examination of PCR products on a 1.7% agarose gel. 1- RT-PCR1, 2- PCR2.

### Step 3: Library cleanup and MiSeq loading

Pool all wells of PCR2 by collecting 4 µL of each.

1. Dilute the pooled library 1:3 (sample : PCR-grade water).

   Note: we find that reducing the dsDNA concentration at this stage increases cleanup quality of the ProNex.
2. Purify the diluted pool with ProNex kit at x1.4 (50 µL sample + 70 µL beads) as follows:
   i. Mix 70 µL ProNex beads (Must be at room temperature before use) and 50 µL sample and incubate for 10 min and place on a magnetic stand for 2 minutes.
   ii. Discarded the supernatant and wash the resin twice by adding 200µl of Wash Buffer for 1 minute and then discarding.
   iii. Allow resin to air-dry for 5 minutes.
   iv. Remove from the magnetic stand and resuspend with 50 µL elution buffer for 5 minutes.
   v. Place the sample on the magnetic stand for 1 minute.
   vi. Transfer the eluted purified HiSpike library to a new tube.
3. Measure the dsDNA concentration of the purified library using DeNovix fluorescence quantification (or an equivalent method).

   Optional: Examine the profile of the purified library using Fragment Analyzer (Figure 2).
4. determine the purified HiSpike library concentration in nM as follws:

$$\frac{C\,\frac{ng}{\mu L}}{649\,\frac{g}{mol} \times Size\ (bp)} \times 10^6 = Molarity\ (nM)$$

5. Dilute the library to 4 nM and confirm using DeNovix.
6. Perform denaturation according to Illumina instruction. Use a final concentration of 12 pM for the run.

7. Use the sample sheet (Supplementary file S5) as the basis for your MiSeq run. This is critical, because of the self-defined indexes used in the method. Copy and paste the names of your samples to the proper field. If you are using different or additional index primers change accordingly their index fields. Update the experiment name and date and remove unused lines.

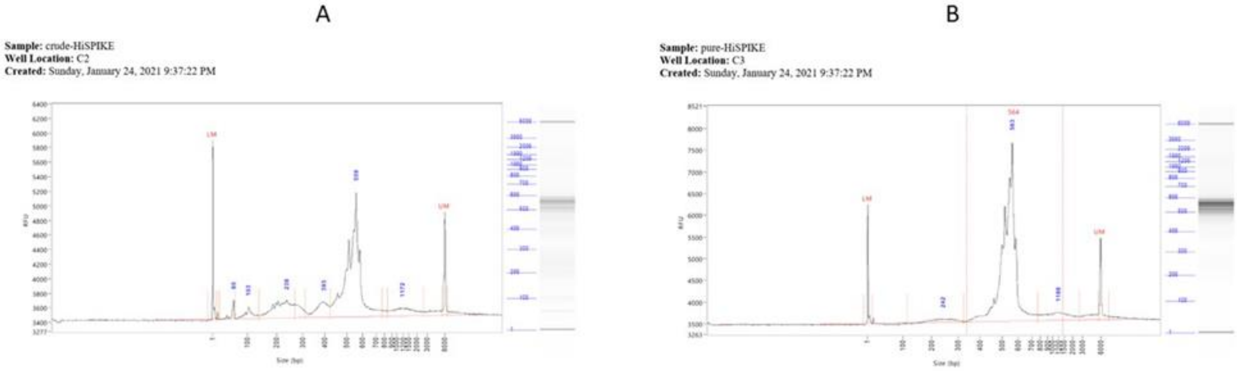


**Figure 2.** Analysis of HiSpike libraries using Fragment Analyzer System (Agilent) with HS NGS Fragment Kit (1-6000bp). **A.** Crude library, **B.** ProNex purified library.

*Bioinformatics*

Sequence acquisition from the Illumina instrument and subsequent processing and analysis can be performed in the routine manner used by your laboratory. We suggest uploading the HiSpike generated spike gene sequences to Nextclade for visualization of clade membership distribution and output of all established and new spike gene mutations in nucleic acid and amino acid notations.

Additional questions concerning setting up and running HiSpike can be addressed to:

Dr. Israel Nissan,

Dr. Ephraim Fass,
